# OncoRAG: Graph-Based Retrieval Enabling Clinical Phenotyping from Oncology Notes Using Local Mid-Size Language Models

**DOI:** 10.64898/2026.03.05.26347717

**Authors:** P. Salome, M. Knoll, D. Walz, N. Cogno, A. S. Dedeoglu, A. L. Qi, S. J. Isakoff, A. Abdollahi, R. B. Jimenez, D. S. Bitterman, H. Paganetti, I. Chamseddine

## Abstract

**Introduction:** Manual data extraction from unstructured clinical notes is labor-intensive and impractical for large-scale clinical and research operations. Existing automated approaches typically require large language models, dedicated computational infrastructure, and/or task-specific fine-tuning that depends on curated data. The objective of this study is to enable accurate extraction with smaller locally deployed models using a disease-site specific pipeline and prompt configuration that are optimized and reusable.

**Materials/Methods:** We developed OncoRAG, a four-phase pipeline that (1) generates feature-specific search terms via ontology enrichment, (2) constructs a clinical knowledge graph from notes using biomedical named entity recognition, (3) retrieves relevant context using graph-diffusion reranking, and (4) extracts features via structured prompts. We ran OncoRAG using Microsoft Phi-3-medium-instruct (14B parameters), a mid-size language model deployed locally via Ollama. The pipeline was applied to three cohorts: triple-negative breast cancer (TNBC; n_patients_=104, n_features_=42; primary development), recurrent high-grade glioma (RiCi; n_patients_=191, n_features_=19; cross-lingual validation in German), and MIMIC-IV (n_patients_=100, n_features_=10; external testing). Downstream task utility was assessed by comparing survival models for 3-year progression-free survival built from automatically extracted versus manually curated features.

**Results:** The pipeline achieved mean F1 scores of 0.80 ± 0.07 (TNBC; n_patients_=44, n_features_=42), 0.79 ± 0.12 (RiCi; n_patients_=61, n_features_=19), and 0.84 ± 0.06 (MIMIC-IV; n_patients_=100, n_features_=10) on test sets under the automatic configuration. Compared to direct LLM prompting and naive RAG baselines, OncoRAG improved the mean F1-score by 0.19 to 0.22 and 0.17 to 0.19, respectively. Manual configuration refinement further improved the F1-score to 0.83 (TNBC) and 0.81 (RiCi), with no change in MIMIC-IV. Extraction time averaged 1.7–1.9 seconds per feature with the 14B model. Substituting a smaller 3.8B model reduced extraction time by 57%, with a decrease in F1-score (0.03–0.10). For TNBC, the extraction time was reduced from approximately two weeks of manual abstraction to under 2.5 hours. In an exploratory survival analysis, models using automatically extracted features showed a comparable C-index to those with manual curation (0.77 vs 0.76; 12 events).

**Conclusions:** OncoRAG, deployed locally using a mid-size language model, achieved accurate feature extraction from multilingual oncology notes without fine-tuning. It was validated against manual extraction for both retrieval accuracy and survival model development. This locally deployable approach, which requires no external data sharing, addresses a critical bottleneck in scalable oncology research.

**Graphical abstract:** 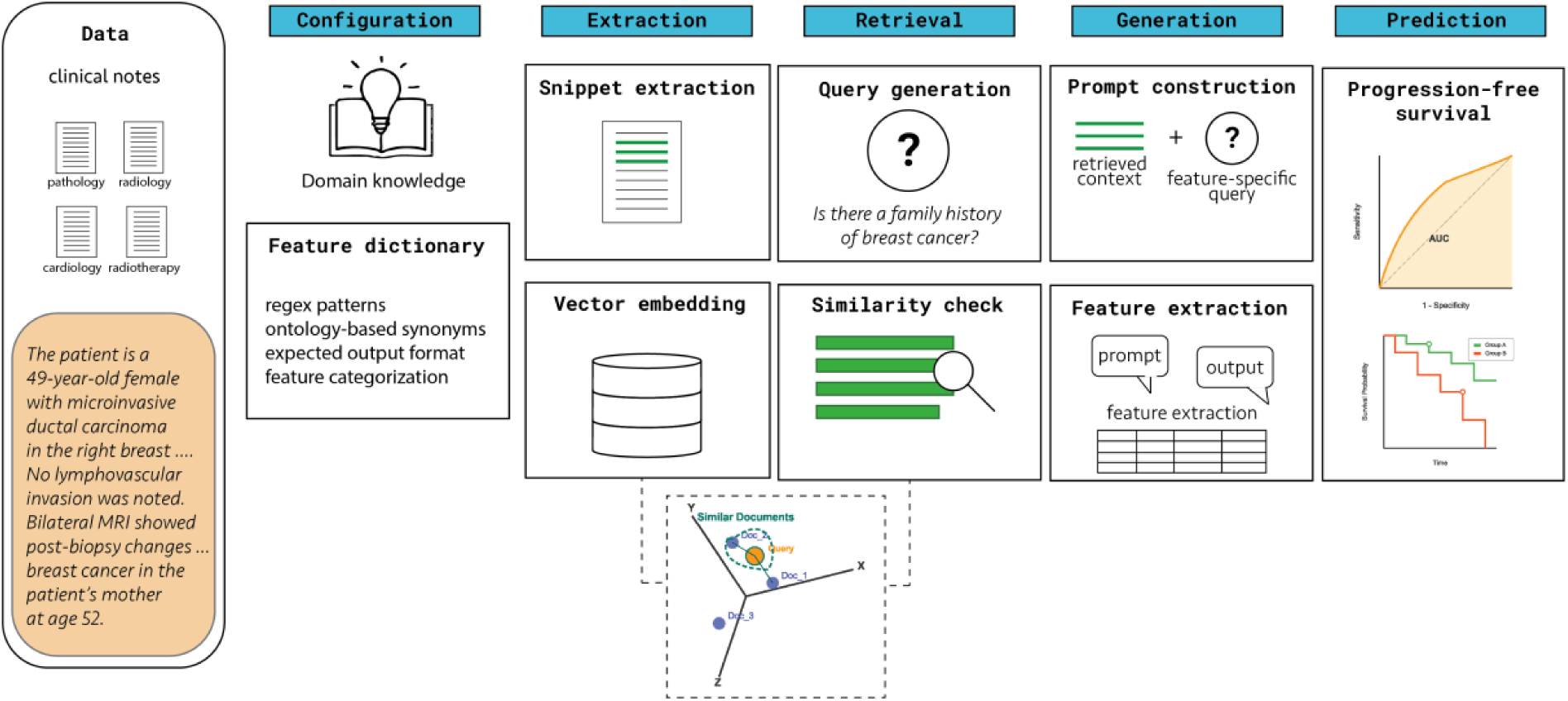

## 1. Introduction

Despite advances in modern cancer care, treatment outcomes remain highly variable, underscoring a need for personalized therapy guided by real-world data [1]. Electronic health records (EHRs) contain vast oncology data that could enable such personalization and accelerate clinical research [2]. Yet, harnessing these data is not straightforward, as EHR data exist in both structured (such as laboratory results, patient demographics) and unstructured (such as clinical notes) formats. Structured data are readily retrievable but lack the granular detail essential for comprehensive oncology research. Unstructured data, including consultation reports, progress notes, and pathology findings, captures clinical details that structured fields cannot represent [3], [4]. This bottleneck is exemplified in radiation oncology; for instance, many features critical for assessing prognosis, such as treatment response or specific toxicities, are documented almost exclusively within clinical narratives [5]. Manual chart abstraction, while accurate, is labor-intensive and impractical for large-scale studies, making automated methods essential for advancing digital oncology [6]. Early efforts to automate this process relied on rule-based systems, but were constrained by hand-crafted rules that could not generalize beyond predefined patterns [7]. Machine learning and transformer models, such as Bidirectional Encoder Representations from Transformers (BERT), overcame this by learning patterns from clinical text, though they required manually labeled training data to perform well [8]. This has led to a growing body of work applying Large Language Models (LLMs) to extract clinical information from oncology text [9], [10], including radiotherapy treatment parameters in radiation oncology [11]. While LLMs have enabled few-shot learning, reducing annotation requirements, challenges remain, including limited context windows and difficulties with factual grounding [12], [13].

Retrieval-Augmented Generation (RAG) offers a promising approach to address these limitations by grounding LLM outputs in retrieved source documents [14]. In its traditional form, RAG retrieves semantically similar text chunks, but this approach struggles to capture relationships between clinical entities or temporal sequences, which are critical for accurate clinical feature extraction in oncology [15]. Several strategies have since emerged to enhance retrieval. The Clinical Entity Augmented Retrieval (CLEAR) framework addressed this by using named entity recognition to identify medical concepts and ontological augmentation to locate relevant passages [16]. Prompt engineering strategies have also contributed to improved LLM performance by using structured prompts that incorporate entity definitions and clinical guidelines [17]. Recent open-source pipelines and fine-tuned LLMs have achieved high accuracy on targeted extraction tasks such as TNM staging [18], [19], while graph-based RAG approaches have shown promise in medical question answering [20]. Existing methods typically require large models (70B+ parameters) with dedicated computational infrastructure or task-specific fine-tuning on curated data to achieve high performance, increasing development burden. Additionally, most validations focus on English-language notes and assess extraction accuracy without demonstrating generalization across institutions or utility for downstream clinical applications such as prognostic modeling.

Here, we present a graph-based RAG pipeline for oncology feature extraction using a locally deployed mid-size open-source language model without task-specific fine-tuning. The pipeline integrates automated ontology enrichment, entity-augmented retrieval, and structured prompt engineering, designed to extract relevant clinical context from noisy narratives and reduce hallucination-related errors. By representing clinical narratives as knowledge graphs, our approach leverages entity co-occurrence and temporal structure to retrieve context that conventional methods often miss. We validated the pipeline in two languages (English and German), two institutional cohorts, and one external public cohort (MIMIC-IV) [21]. Extraction accuracy was assessed against manual curation, and practical utility was evaluated by comparing the performance of prognostic models built from automated versus manual data.

## 2. Methods

### 2.1 Study Design and Data Splits

The system was developed and evaluated using three cohorts (Figure 1). The TNBC cohort was split temporally into a development set (n=60; 58%; 2019–2023) and a held-out test set (n=44; 42%; 2023–2024). The RiCi cohort was similarly split temporally into a development set (n=130; 68%; German; 2009–2015) and a held-out test set (n=61; 32%; 2015–2019). Pipeline architecture development and tuning were performed exclusively on the TNBC development set, after which the architecture was locked. Feature configurations (keywords, synonyms, feature definitions, and prompt rules) for each clinical variable were developed on each cohort’s development set. Two configuration approaches were evaluated: an automatic configuration, generated from feature names and expected output categories, and a hybrid configuration, in which automatic settings were manually refined by reviewing extraction errors on the development set until feature-level F1 reached ≥ 0.85. For MIMIC-IV (n=100), 10 features overlapping with the TNBC feature list were identified. Because configurations for these features had already been established during TNBC development, they were applied directly without cohort-specific adaptation, serving as external testing (i.e., no MIMIC-specific configuration or tuning was performed). After all configurations were fixed, independent testing was conducted on the held-out TNBC test set, the held-out RiCi test set, and the full MIMIC-IV cohort.

**Figure 1.**
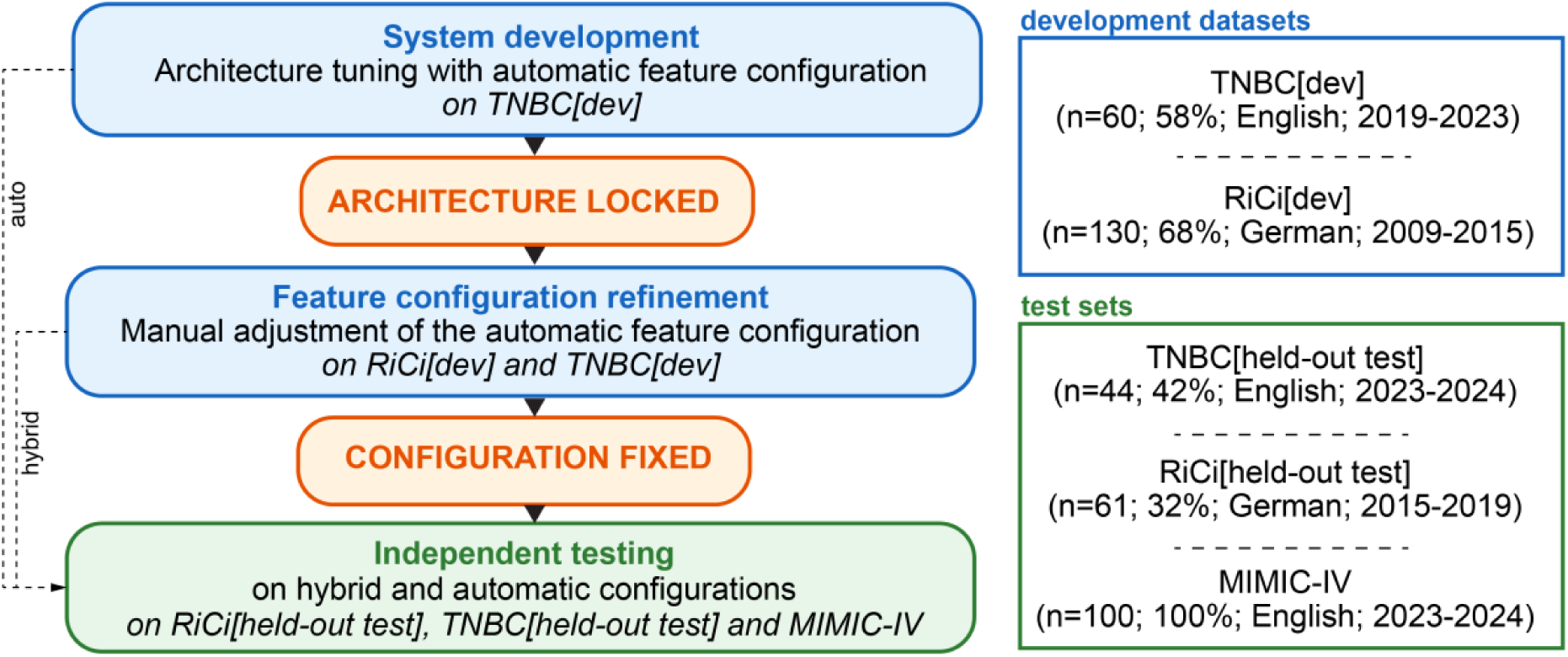
Study design and data splits. System development and architecture tuning were performed on the TNBC development set (Step 1), after which the pipeline architecture was locked. Feature configurations were then developed on each cohort’s development set; the hybrid configuration was further refined by reviewing errors on development data only (Step 2). Independent testing (Step 3) was conducted on temporally held-out patients and the independent test set MIMIC-IV. Blue indicates development phases; green indicates testing phases; orange indicates checkpoints; auto: automatic.

### 2.2 Study Population and Data Sources

Across all three cohorts, only unstructured clinical notes were used as input to the pipeline; structured EHR fields, such as laboratory results, coded diagnoses, and demographic tables, were not included in the extraction process.

#### 2.2.1 Triple Negative Breast Cancer Cohort

The pipeline was developed and tested on a retrospective cohort of 104 female patients with triple-negative breast cancer (TNBC) treated at Massachusetts General Hospital between 2019 and 2024. Patients received neoadjuvant systemic therapy (chemotherapy and/or immunotherapy) followed by surgery, with adjuvant systemic or radiation therapy administered in select cases. Clinical notes were extracted from the Epic electronic health record system, including surgical reports, pathology reports, imaging reports, oncology consultations, radiation oncology notes, and discharge summaries, totaling 51,099 pages (mean: 490 pages per patient). Median follow-up was 18 months (interquartile range [IQR]: 15-21 months).

#### 2.2.2 Recurrent High-Grade Glioma Cohort

To evaluate cross-lingual and cross-institutional performance, we used a cohort of 191 patients with recurrent high-grade glioma treated with carbon ion re-irradiation (RiCi) at the Heidelberg Ion Beam Therapy Center (Germany, 2009-2018) [22], [23]. All patients had received prior standard-of-care treatment. Clinical notes in German included physician letters, pathology reports, surgical reports, imaging reports, and tumor board discussions, totaling 3,076 pages (mean: 16 pages per patient). Median follow-up was 10 months (IQR: 6-14 months).

#### 2.2.3 MIMIC-IV Cohort

This cohort included a subset of the publicly available MIMIC-IV (Medical Information Mart for Intensive Care) dataset, which contains de-identified ICU notes from Beth Israel Deaconess Medical Center (2008–2019) [21], [24]. The subset includes 100 ICU patients with structured fields that served as ground truth for validating 10 clinical features overlapping with the TNBC feature list [25]. Feature inclusion was restricted to variables whose corresponding information was documented in the unstructured clinical notes in at least 70% of patients, ensuring that the ground truth derived from structured fields could be compared against note-based extraction. Notes included discharge summaries, radiology reports, nursing progress notes, and specialist consults, totaling 5,646 pages (mean: 56 pages per patient). Because these features utilized the configuration files already established during the TNBC phase, no development/test split was required; the entire cohort served as a testing set to evaluate both the automatic and manually adjusted configurations.

#### 2.2.4 Ground Truth Annotation

For the TNBC cohort, variables were manually extracted by a trained researcher using a pre-defined data dictionary specifying clinical definitions, allowed categorical values, and a prioritization hierarchy for conflicting documentation based on the American Joint Committee on Cancer (AJCC) [26] and the North American Association of Central Cancer Registries (NAACCR) standards [27], which favor high-specificity sources like pathology and surgical reports over clinical notes for tumor staging and biomarker status. The complete extraction was performed three times at two-week intervals, and discrepancies and difficult cases were resolved by an attending radiation oncologist; unrecorded values were marked as missing. For the RiCi cohort, ground truth values had been previously curated by two annotators and validated in prior studies [23]. For the MIMIC-IV cohort, ground truth was derived from structured database fields; validation was limited to 10 features for which corresponding fields existed, with no patient-level exclusions.

#### 2.2.5 Ethical Approval

For the TNBC cohort from Massachusetts General Brigham, this retrospective study was conducted in compliance with the Health Insurance Portability and Accountability Act and approved by the Institutional Review Board (Protocol #2024P001234) with waiver of informed consent. The recurrent glioma cohort analysis was conducted in accordance with the Declaration of Helsinki and approved by the Institutional Review Board of the Medical Faculty of Heidelberg University (approval number S-540/2010; last updated 20 July 2020). MIMIC-IV data access was obtained from PhysioNet after completing the required data use agreements and CITI training.

### 2.3 Feature Selection and Definition

Clinical features were selected for extraction based on their relevance to clinical outcomes in each disease site, as well as availability in clinical notes. For TNBC, 42 features were defined, spanning patient demographics, clinical history, tumor characteristics, biomarkers, treatment details, and outcomes [28]. For RiCi, 19 features were selected based on variables previously analyzed in studies of recurrent high-grade glioma [23]. For MIMIC-IV external validation, 10 features that overlapped with the TNBC configuration and had available ground truth in MIMIC-IV structured fields were used. Features were classified as binary categorical (e.g., diabetes: yes/no), multiclass categorical (e.g., tumor quadrant; clock position), or originally continuous variables that were binarized for evaluation (pack-years: 0 vs >0; parity: 0 vs ≥1). Missing data were handled differently by variable type: for binary categorical variables, if a condition or treatment was not mentioned, then we assumed that the patient did not have that condition or receive that treatment; for multiclass categorical and continuous variables, undocumented values were kept as missing rather than imputed. For biomarker features where testing may not have been performed (IDH1 mutation, MGMT methylation, 1p/19q co-deletion), undocumented values were retained as a separate missing category rather than assumed absent, as the absence of documentation reflects an untested or unrecorded status rather than a negative result. The complete feature lists are provided in Supplementary Table S1.

### 2.4 The OncoRAG Pipeline Architecture

The automated extraction pipeline has four sequential phases (Figure 2). Medical ontologies used for feature enrichment were Unified Medical Language System (UMLS) [29] and BioPortal [30]. All pipeline parameters were tuned using the TNBC development set. The following sections describe each phase; additional implementation details are provided in the Supplementary Methods.

**Figure 2.**
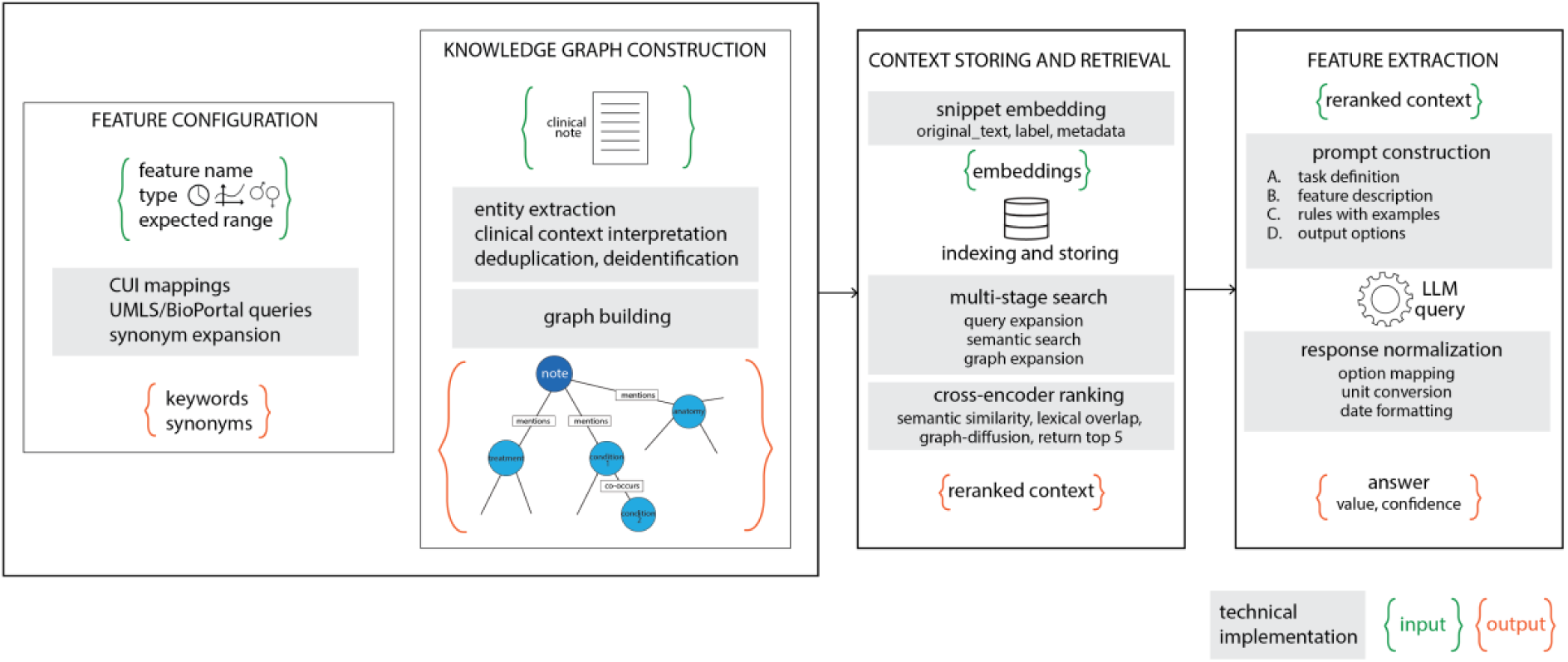
The automated feature extraction pipeline for generating structured TNBC clinical features. The process consists of four phases: (1) Configuration generation, where features, ontologies, and extraction rules/queries are defined; (2) Knowledge graph construction, which involves biomedical named entity recognition, and organization of extracted entities into a clinical knowledge graph; (3) Context retrieval, utilizing entity indexing, semantic search, and graph-diffusion reranking to select relevant context sentences; and (4) Feature extraction, where structured prompts with retrieved context are submitted to the language model for value extraction and post-processing to produce the final structured data.

#### 2.4.1 Phase 1: Automated Feature Configuration

Each feature was defined by a set of attributes: name, data type (categorical or continuous), expected value categories, and whether it was a demographic feature. Non-demographic features underwent automated ontological enrichment via UMLS [29] and BioPortal [30] API queries to generate Concept Unique Identifiers (CUIs) and synonyms (top 5 results per feature, with relevance scores > 0.6), while demographic features were enriched using the language model. Keywords were expanded from feature names and ontology terms using WordNet [31], producing a list of expanded keywords and synonyms for each variable.

#### 2.4.2 Phase 2: Clinical Knowledge Graph Construction

Clinical entities, such as diseases, medications, procedures, and anatomical structures, were extracted using multiple biomedical named entity recognition (NER) models, including both English- and German-language models to support multilingual extraction [32], [33], [34]. Context modifiers, including negation, temporality, family history, and hypothetical status, were detected using medspaCy [35]. Entity deduplication employed SapBERT embeddings to cluster semantically similar entities (cosine similarity ≥0.85). Knowledge graphs were then constructed using NetworkX [36] (Figure S1; Supplementary Table S2) with nodes representing patient identifiers, notes, dates, and the resulting entities, mapped to standardized categories (Condition, Treatment, Procedure, Anatomy, Gene, Protein). Edges linked patients to their notes, notes to mentioned entities (preserving the source sentence for each mention), entities to dates, and co-occurring entities to each other. Relationships between entities were captured through co-occurrence within clinical notes rather than explicit relation extraction.

#### 2.4.3 Phase 3: Entity Indexing and Context Retrieval

Entity nodes from the knowledge graph were indexed in ChromaDB using SapBERT embeddings [37]. Retrieval proceeded through multiple stages: query expansion generated up to 80 search terms (median approximately 60) from feature keywords and ontology synonyms; semantic search retrieved the top 30 matching entities; and structural expansion via breadth-first search (2 hops) captured related nodes, with edges weighted by relationship type. Candidate entities were ranked by relevance scores weighted by entity type, and the top 30 were selected.

Because semantic retrieval alone can return irrelevant context, reranking was applied to prioritize informative sentences. Context sentences were scored with multiple signals: semantic similarity from the BioBERT-NLI cross-encoder [38] (S_sem_); lexical overlap (S_lex_) as a weighted term match over feature keywords, synonyms, and expected values; name/synonym matching (S_name_) via additional cross-encoder queries using the same ontology-enriched term list; and a binary keyword boost (B_kw_) applied when any boosted term was present. A graph-diffusion reranker (S_graph_) further refined results by constructing a cosine-similarity graph over candidate sentences and smoothing embeddings across neighbors. Hypothetical and planning mentions were filtered, while negated, family, and historical evidence were retained for downstream interpretation. The final retrieval score, used to select the top 5 sentences for feature extraction, was computed as:

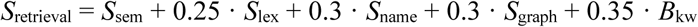

#### 2.4.4 Phase 4: LLM-Based Feature Extraction

Feature values were extracted from the top 5 retrieved sentences using structured prompts submitted to Microsoft Phi models (Phi-3.5-mini-instruct [3.8B parameters] and Phi-3-medium-instruct [14B parameters]) [39], which were run locally via Ollama. For time-varying features, temporal anchoring was applied: baseline features used the value closest to the initial cancer diagnosis (within ±9 months) for the TNBC and RiCi cohorts and the first reported value for MIMIC-IV, while outcome features used the last documented value. The same temporal criteria were applied during manual ground truth curation to ensure comparability. Prompts contained four components: task definition, feature description, interpretation rules with examples, and multiple-choice output options (Supplementary Table S3). Feature descriptions and examples were generated using the LLM and enriched with ontological and lexical sources. For categorical features, models returned a single letter corresponding to the correct option; for numeric and date features, values were normalized (unit conversion and date standardization where applicable). For each extraction, the LLM reported a self-assessed confidence level (High, Medium, or Low) based on the clarity of supporting evidence. A confidence-weighted retrieval score (S_retrieval_×w_conf_, where w_conf_ = 1.0/0.7/0.4 for High/Medium/Low) was computed to assess the association between retrieval quality and extraction accuracy. All experiments were conducted on a local workstation equipped with an NVIDIA RTX A5500 GPU (24 GB VRAM), 32 CPU cores, and 126 GB RAM.

### 2.5 Extraction Accuracy Evaluation

Four experiments evaluated pipeline components and extraction accuracy. First, OncoRAG was compared against two baselines: direct LLM prompting without retrieval (LLM-only), in which the same structured prompts were submitted to the model without any retrieved context, and naive vector-based RAG, in which the top 5 semantically similar text chunks were retrieved using cosine similarity over SapBERT embeddings without graph construction or reranking. Second, we assessed the impact of context reranking by comparing feature extraction using raw semantic retrieval (top 5 sentences) against the full reranking pipeline. Third, model architecture tested both Phi models (Phi-3-medium [14B] and Phi-3.5-mini [3.8B]) with 4K and 128K context windows, yielding four experimental conditions. Fourth, configuration quality was compared between automated configurations (LLM classification and ontology enrichment) and manually refined versions. Evaluation used exact-match scoring against manual ground truth, with precision, recall, F1-score, and accuracy computed per class and macro-averaged. Statistical comparisons of confidence-weighted retrieval scores between correct and incorrect predictions were computed at the patient level and compared using Mann-Whitney U tests.

### 2.6 Downstream Task Utility: Prognostic Modeling

Prognostic modeling, especially using machine learning, is an active research area hindered by a lack of structured data, which prevents large-scale feature selection and the evaluation of new biomarkers and their interactions. To evaluate the potential of OncoRAG to address this limitation, we fit two ridge-penalized Cox proportional hazards models for 3-year progression-free survival in the TNBC cohort (n=101; 3 patients excluded due to insufficient follow-up; 12 events): one using automatically extracted features and one using manually curated features. Given the low event count, this analysis is exploratory and intended to assess whether automatically extracted features preserve sufficient signal for prognostic modeling. Features with at least 20% missing data were excluded; remaining missing values were median-imputed, and collinear features (Spearman’s ρ > 0.80) were removed. Inputs were standardized (zero mean, unit variance) and reduced via principal component analysis (PCA) to 5 components (approximately 67% variance explained). Model performance was evaluated using the concordance index (C-index) with 1000-sample bootstrap estimation. Survival curves were generated comparing the mean model-predicted survival with 95% bootstrap confidence bands.

## 3. Results

### 3.1 Feature Extraction Performance

Table 1 summarizes macro-averaged precision, recall, F1-score, and accuracy aggregated across all extracted features for the TNBC, RiCi, and MIMIC-IV cohorts. On independent test sets, the automatic configuration achieved mean precision, recall, F1-score, and accuracy of 0.79, 0.87, 0.80, and 0.85 (TNBC, 42 features), 0.80, 0.83, 0.79, and 0.87 (RiCi, 19 features), and 0.84, 0.88, 0.84, and 0.90 (MIMIC-IV, 10 features). Compared to direct prompting without retrieval (LLM-only), OncoRAG improved the mean F1-score by 0.20, 0.22, and 0.19 in TNBC, RiCi, and MIMIC-IV, respectively. Compared to naive vector-based RAG, improvements were 0.19, 0.17, and 0.17. The full reranking pipeline improved mean F1-score by 0.10 compared to raw semantic retrieval on the TNBC development set (0.81 vs 0.71), 0.09 on the RiCi development set (0.82 vs 0.74), and 0.04 on MIMIC-IV (0.84 vs 0.80). The hybrid configuration further improved precision, recall, F1-score, and accuracy by 0.02, 0.02, 0.03, and 0.02 in TNBC and 0.04, 0.00, 0.02, and 0.02 in RiCi, with no change in MIMIC-IV.

**Table 1.**
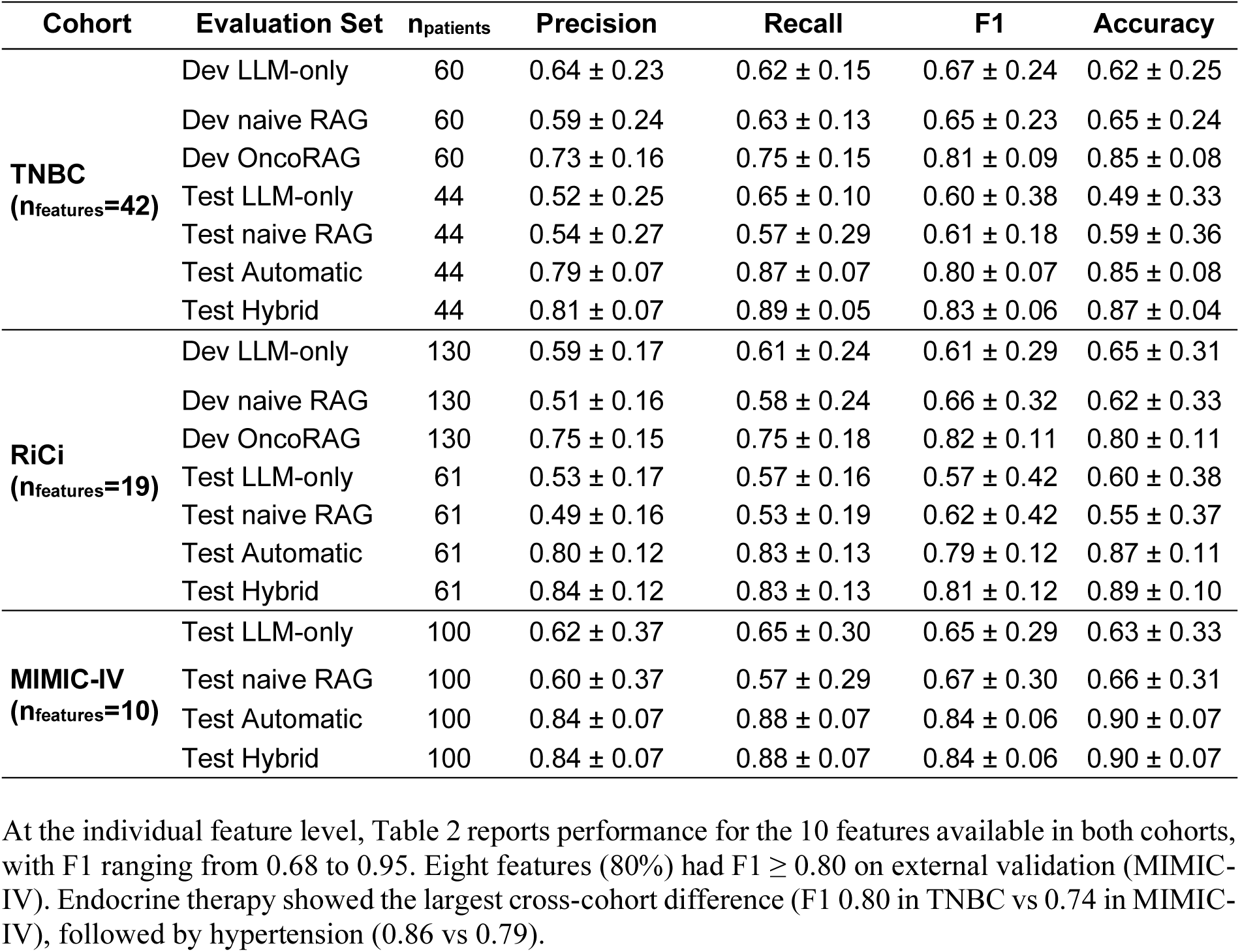
Feature extraction performance by cohort and method. Values represent macro-averaged precision, recall, F1-score, and accuracy (mean ± SD across features). Baseline methods were evaluated on both development (Dev) and held-out test sets; OncoRAG results are reported under two configurations: automatic (generated solely from feature names and ontology enrichment) and hybrid (automatic configuration manually refined on the development set). For MIMIC-IV, TNBC-derived configurations were applied without cohort-specific adaptation, and the entire cohort served as a test set. LLM-only, direct prompting without retrieval; naive RAG, vector-based retrieval-augmented generation; OncoRAG, graph-based retrieval-augmented generation. TNBC, triple-negative breast cancer; RiCi, recurrent high-grade glioma; n_patients_, number of patients; n_features_, number of features.

| Cohort | Evaluation Set | $n_{patients}$ | Precision | Recall | F1 | Accuracy |
| --- | --- | --- | --- | --- | --- | --- |
| **TNBC**<br>($n_{features}=42$) | Dev LLM-only | 60 | 0.64 ± 0.23 | 0.62 ± 0.15 | 0.67 ± 0.24 | 0.62 ± 0.25 |
| | Dev naive RAG | 60 | 0.59 ± 0.24 | 0.63 ± 0.13 | 0.65 ± 0.23 | 0.65 ± 0.24 |
| | Dev OncoRAG | 60 | 0.73 ± 0.16 | 0.75 ± 0.15 | 0.81 ± 0.09 | 0.85 ± 0.08 |
| | Test LLM-only | 44 | 0.52 ± 0.25 | 0.65 ± 0.10 | 0.60 ± 0.38 | 0.49 ± 0.33 |
| | Test naive RAG | 44 | 0.54 ± 0.27 | 0.57 ± 0.29 | 0.61 ± 0.18 | 0.59 ± 0.36 |
| | Test Automatic | 44 | 0.79 ± 0.07 | 0.87 ± 0.07 | 0.80 ± 0.07 | 0.85 ± 0.08 |
| | Test Hybrid | 44 | 0.81 ± 0.07 | 0.89 ± 0.05 | 0.83 ± 0.06 | 0.87 ± 0.04 |
| **RiCi**<br>($n_{features}=19$) | Dev LLM-only | 130 | 0.59 ± 0.17 | 0.61 ± 0.24 | 0.61 ± 0.29 | 0.65 ± 0.31 |
| | Dev naive RAG | 130 | 0.51 ± 0.16 | 0.58 ± 0.24 | 0.66 ± 0.32 | 0.62 ± 0.33 |
| | Dev OncoRAG | 130 | 0.75 ± 0.15 | 0.75 ± 0.18 | 0.82 ± 0.11 | 0.80 ± 0.11 |
| | Test LLM-only | 61 | 0.53 ± 0.17 | 0.57 ± 0.16 | 0.57 ± 0.42 | 0.60 ± 0.38 |
| | Test naive RAG | 61 | 0.49 ± 0.16 | 0.53 ± 0.19 | 0.62 ± 0.42 | 0.55 ± 0.37 |
| | Test Automatic | 61 | 0.80 ± 0.12 | 0.83 ± 0.13 | 0.79 ± 0.12 | 0.87 ± 0.11 |
| | Test Hybrid | 61 | 0.84 ± 0.12 | 0.83 ± 0.13 | 0.81 ± 0.12 | 0.89 ± 0.10 |
| **MIMIC-IV**<br>($n_{features}=10$) | Test LLM-only | 100 | 0.62 ± 0.37 | 0.65 ± 0.30 | 0.65 ± 0.29 | 0.63 ± 0.33 |
| | Test naive RAG | 100 | 0.60 ± 0.37 | 0.57 ± 0.29 | 0.67 ± 0.30 | 0.66 ± 0.31 |
| | Test Automatic | 100 | 0.84 ± 0.07 | 0.88 ± 0.07 | 0.84 ± 0.06 | 0.90 ± 0.07 |
| | Test Hybrid | 100 | 0.84 ± 0.07 | 0.88 ± 0.07 | 0.84 ± 0.06 | 0.90 ± 0.07 |

At the individual feature level, Table 2 reports performance for the 10 features available in both cohorts, with F1 ranging from 0.68 to 0.95. Eight features (80%) had F1 ≥ 0.80 on external validation (MIMIC-IV). Endocrine therapy showed the largest cross-cohort difference (F1 0.80 in TNBC vs 0.74 in MIMIC-IV), followed by hypertension (0.86 vs 0.79).

**Table 2.** Cross-cohort generalizability of OncoRAG feature extraction for the 10 overlapping clinical variables under the automatic configuration. Values represent macro-averaged precision, recall, and F1-score (mean ± SD across features), computed per class and averaged across all classes for each feature. Accuracy represents the proportion of correctly classified instances. Class Dist., class distribution within each cohort’s test set. TNBC, triple-negative breast cancer; MIMIC-IV, Medical Information Mart for Intensive Care IV; HRT, hormone replacement therapy; Pre, premenopausal; Post, postmenopausal; N, no; Y, yes; n, number of patients.

| TNBC (n=44) |  |  |  |  |  | MIMIC-IV (n=100) |  |  |  |  |
| --- | --- | --- | --- | --- | --- | --- | --- | --- | --- | --- |
| Feature | Class Dist. | P | R | F1 | Acc | Class Dist. | P | R | F1 | Acc |
| HRT Use | N:37, Y:7 | 0.73 | 0.85 | 0.77 | 0.84 | N:96, Y:4 | 0.87 | 0.87 | 0.87 | 0.98 |
| Menopause | Pre:26, Post:18 | 0.88 | 0.88 | 0.86 | 0.86 | NA:57, Pre:12, Post:31 | 0.83 | 0.82 | 0.82 | 0.85 |
| Parity | 0:11, ≥1:33 | 0.82 | 0.88 | 0.84 | 0.86 | NA:57, 0:26, ≥1:17 | 0.95 | 0.95 | 0.95 | 0.97 |
| Substance Use | N:28, Y:16 | 0.87 | 0.88 | 0.88 | 0.89 | N:81, Y:19 | 0.91 | 0.93 | 0.92 | 0.95 |
| Diabetes | N:38, Y:6 | 0.80 | 0.95 | 0.85 | 0.91 | N:64, Y:36 | 0.86 | 0.83 | 0.84 | 0.86 |
| Metformin | N:41, Y:3 | 0.71 | 0.95 | 0.77 | 0.91 | N:89, Y:11 | 0.84 | 0.97 | 0.89 | 0.95 |
| Hypertension | N:16, Y:28 | 0.85 | 0.88 | 0.86 | 0.86 | N:32, Y:68 | 0.79 | 0.84 | 0.79 | 0.80 |
| Smoking | N:33, Y:11 | 0.76 | 0.83 | 0.77 | 0.80 | N:61, Y:39 | 0.80 | 0.81 | 0.80 | 0.81 |
| Alcohol Use | N:38, Y:6 | 0.68 | 0.86 | 0.68 | 0.75 | N:77, Y:23 | 0.90 | 0.75 | 0.80 | 0.88 |
| Endocrine Tx | N:33, Y:11 | 0.79 | 0.83 | 0.80 | 0.84 | N:98, Y:2 | 0.67 | 0.98 | 0.74 | 0.96 |

Figure 3 presents macro-averaged F1-scores across development and test sets. In TNBC, 27 of 42 features (64%) achieved F1 ≥ 0.80 on the hybrid test set. In RiCi, 12 of 19 features (63%) achieved F1 ≥ 0.80. On MIMIC-IV, 8 of 10 features (80%) achieved F1 ≥ 0.80 (Figure 3A). Complete feature-level results are provided in Supplementary Tables S4–S6.

**Figure 3.**
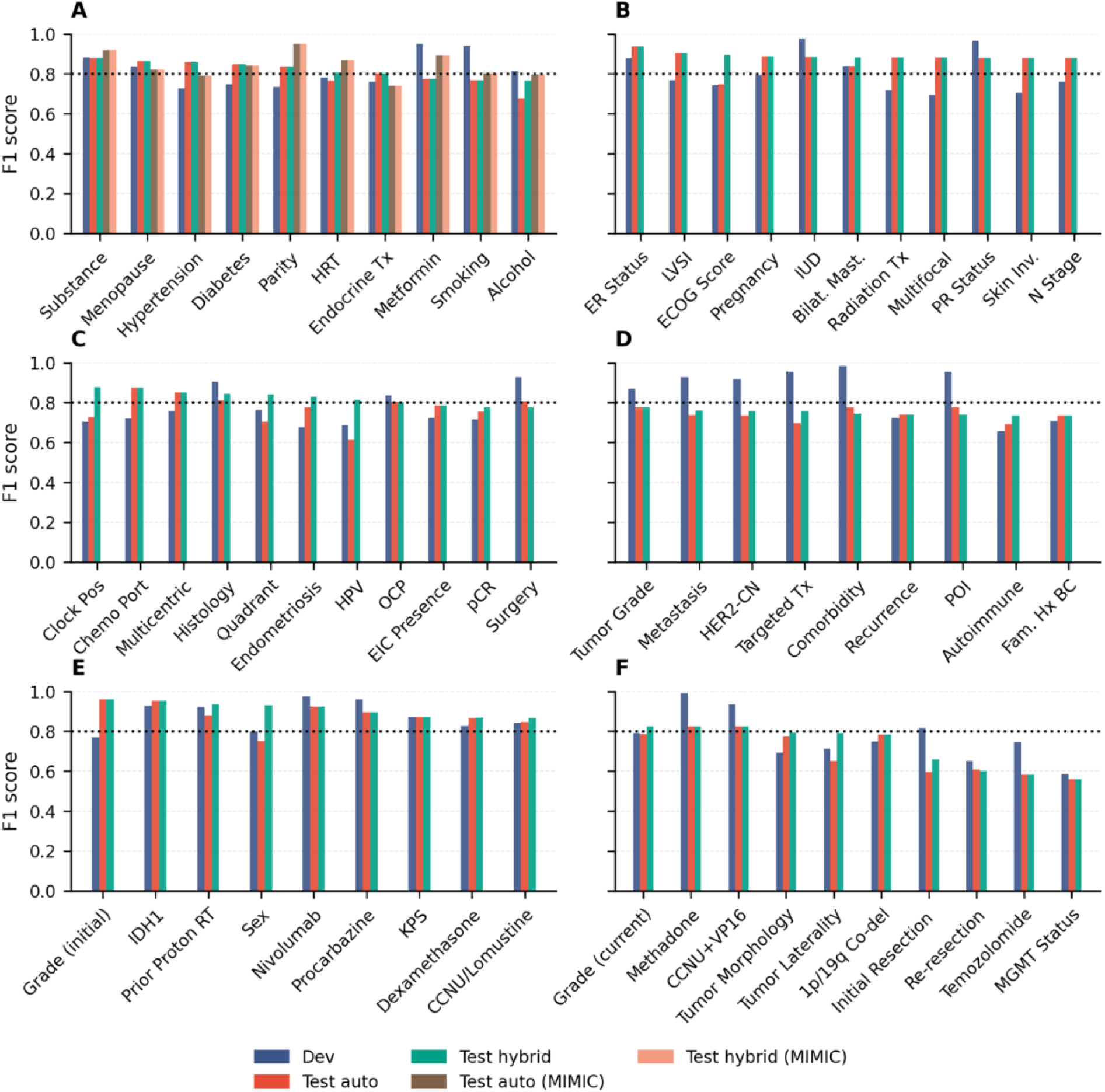
Feature-level macro-averaged F1-scores across development, test automatic, and test hybrid sets. (A) Features with external MIMIC-IV validation (test automatic and test hybrid shown for both TNBC and MIMIC-IV). (B–D) TNBC features are grouped by performance tiers. (E–F) RiCi cohort features. The dashed line indicates the F1 = 0.80 threshold. Features are defined in Supplementary Table S1.

### 3.2 Impact of Model Size and Context Window

Figure 4A shows the performance across models and context window sizes. We evaluated four model configurations varying in size (Phi-3-medium [14B] vs. Phi-3.5-mini [3.8B]) and context window (128k vs. 4k tokens). The medium model with 128k context achieved the highest performance in TNBC (F1 = 0.81) and MIMIC-IV (F1 = 0.84), while the medium model with 4k context performed best in RiCi (F1 = 0.84). Reducing from the medium to the mini model decreased the mean F1-score by 0.10 in TNBC and 0.03 in RiCi. Features most affected included pregnancy (−0.37), hypertension (−0.31), and substance use (−0.28). Context window effects varied by cohort: in RiCi, MGMT status (0.17), sex (0.09), and morphology (0.09) improved with a smaller context, while in TNBC, substance use (−0.18) and ER status (−0.10) performed better with a larger context. Extraction time per feature averaged 1.66–1.91s for Phi-3-medium and 0.72–0.78s for Phi-3.5-mini across cohorts, representing a 57% reduction in processing time. Reducing the context window size (128k to 4k) had a minimal impact on the mean F1-score (difference < 0.02).

**Figure 4.**
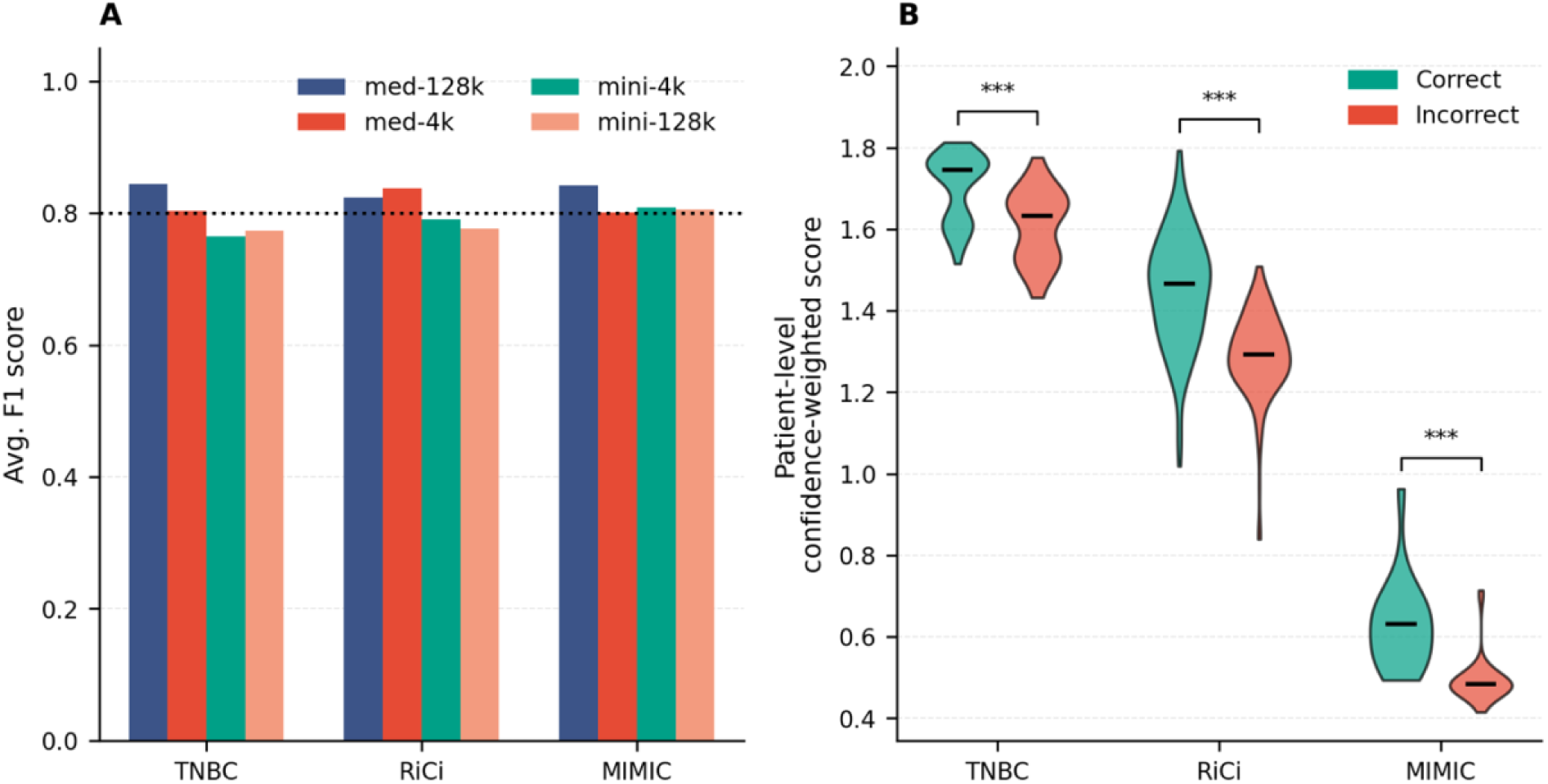
Extraction performance and retrieval quality. (A) Macro-averaged F1-scores across cohorts for medium and mini models with 128k and 4k context windows. The dotted line indicates F1 = 0.80. (B) Patient-level mean confidence-weighted retrieval scores for correct versus incorrect predictions. ***p < 0.001, Mann-Whitney U test.

### 3.3 Error Analysis

In TNBC, lower-performing features (F1 < 0.80) included HPV status (0.61), alcohol use (0.68), autoimmune disease (0.69), targeted therapy (0.70), and quadrant (0.70). In RiCi, MGMT status (0.56), temozolomide use (0.58), initial resection (0.60), re-resection status (0.61), and laterality (0.65) showed lower performance. Retrieval scores ranged from 0.41 to 1.95 across features, with considerable variation by cohort: MIMIC-IV (0.41–1.95), RiCi (0.90–1.95), and TNBC (1.14–1.95). Manual review of misclassified instances indicated that 62% of errors were attributable to retrieval failures (relevant context not in the top 5 sentences) and 38% to LLM interpretation errors despite correct context retrieval. As shown in Figure 4B, correct predictions had significantly higher patient-level mean confidence-weighted retrieval scores than incorrect predictions: TNBC (1.61 vs 1.55), RiCi (1.73 vs 1.60), and MIMIC-IV (0.87 vs 0.61; all p < 0.001, Mann-Whitney U test).

### 3.4 Downstream Task Utility: Prognostic Modeling

For 3-year PFS prediction in the TNBC cohort, bootstrap estimation on the full cohort yielded C-indices of 0.77 (95% CI: 0.64–0.86) for the automatic model and 0.76 (95% CI: 0.65–0.86) for manual curation (p = 0.512, paired bootstrap test; Figure 5). Automatic extraction completed in approximately 2.5 hours compared to two weeks for manual chart abstraction.

**Figure 5.**
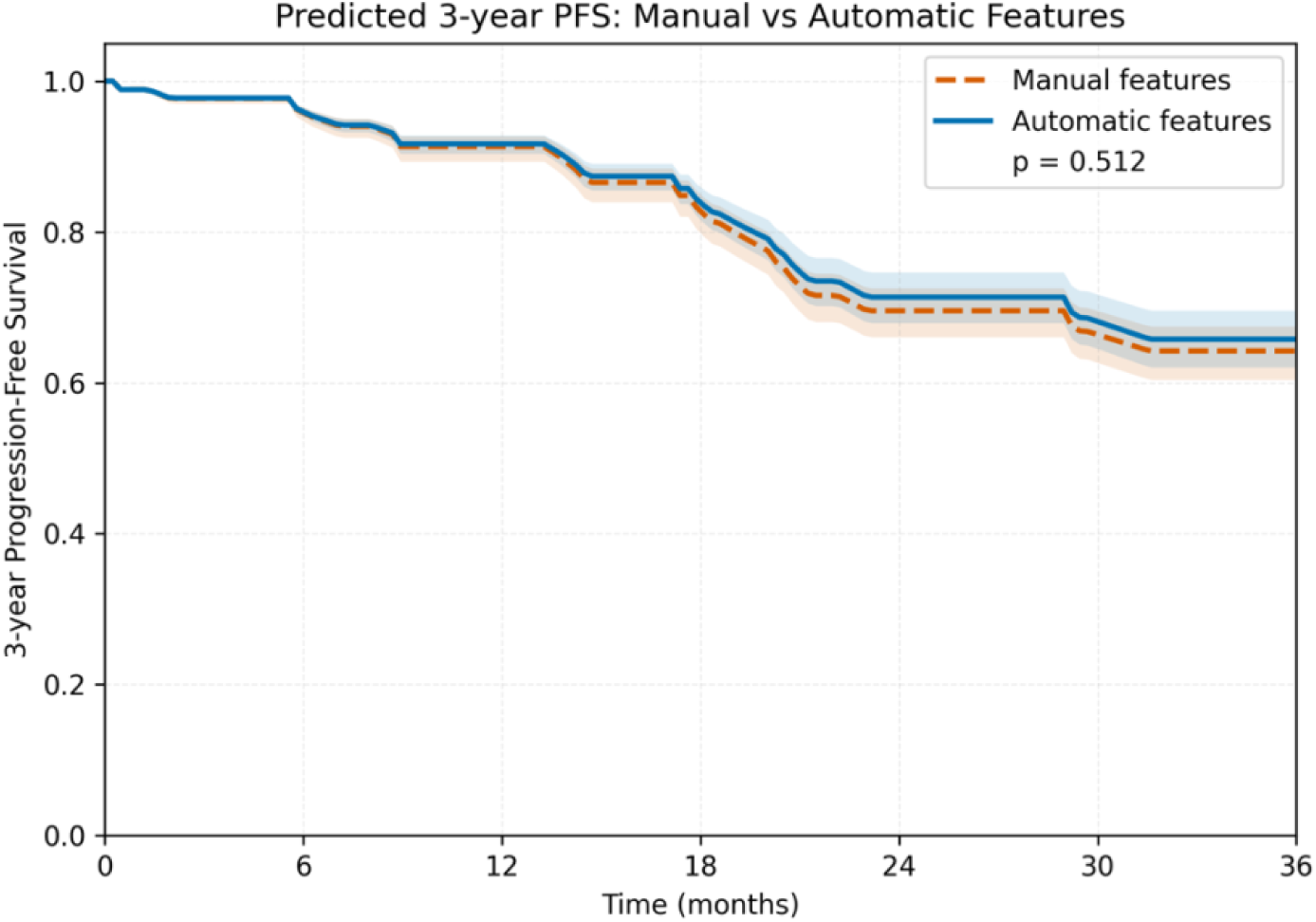
Predicted 3-year progression-free survival comparing models trained on manually curated features (dashed orange) versus automatically extracted features (solid blue). Shaded bands represent 95% bootstrap confidence intervals (1000 resamples). p = 0.512, paired bootstrap test on C-index difference.

## 4. Discussion

Automated extraction of clinical features from unstructured oncology notes has typically relied on rule-based systems, supervised machine learning, or large-scale language models. Rule-based and dictionary approaches require extensive manual curation and lack generalizability across institutions [40], while supervised methods demand large annotated training sets for each target feature [41]. Recent LLM-based approaches achieve broader coverage but often require either large-scale models (70B+ parameters) or extensive fine-tuning to achieve research-grade accuracy [42]. Unlike these methods, OncoRAG does not rely on the language model to reason over raw clinical text; instead, clinical logic is structured through a pre-inference orchestration layer. As a result, a locally deployed mid-size language model (14B parameters) can achieve research-grade accuracy without fine-tuning, yielding survival models with discrimination comparable to that from manual curation.

Our pipeline addresses several limitations of existing methods. By representing clinical narratives as knowledge graphs rather than text chunks, we leverage entity co-occurrence and temporal structure that conventional RAG approaches miss. Compared to direct LLM prompting without retrieval, OncoRAG improved the mean F1-score by 0.19 to 0.22 across cohorts, and by 0.17 to 0.19 compared to naive vector-based RAG (Table 1). The 0.04 to 0.10 F1-score improvement from graph-diffusion reranking over raw semantic retrieval confirms that structured context selection is essential for extraction accuracy. This is enabled by our use of specialized biomedical NER models for graph construction, reserving LLM inference solely for final feature extraction, reducing computational overhead while maintaining accuracy. The hybrid configuration provided modest gains over automatic configuration in institutional cohorts (0.02–0.03 F1); MIMIC-IV showed no difference, primarily because it was evaluated using TNBC-derived configurations without cohort-specific refinement. This suggests that the benefits of hybrid refinement are cohort-specific and do not transfer across institutions.

Error analysis indicated that 62% of failures originated from retrieval rather than LLM interpretation, suggesting that further gains require improved retrieval strategies rather than larger models. Across cohorts, the pipeline exhibited higher recall than precision (Table 1), indicating a tendency to overidentify rather than miss features. Higher recall than precision is expected given the class imbalance in clinical datasets and may represent a favorable trade-off for downstream use: false positives are easier to identify and correct than false negatives, which may go undetected. Lower-performing features fell into three categories: social history variables with conflicting documentation, tumor characteristics better captured by imaging-based approaches (such as tumor quadrant and clock position), and features with sparse documentation, ambiguous terminology, or complex reasoning across multiple reports.

Validation across two languages, three institutions, and two cancer types demonstrates generalizability beyond single-center, English-only datasets that characterize most prior work. The 57% reduction in extraction time with the smaller model, with minimal accuracy loss (F1 difference < 0.02), enables locally deployable extraction suitable for sensitive clinical data without cloud-based inference. From a clinical research perspective, this is directly relevant to a persistent bottleneck in the field: the labor-intensive chart abstraction required to link imaging-derived features with clinical phenotypes for outcome prediction studies [43].

This study has several limitations. Manual adjustments in the hybrid configuration require domain expertise, which may constrain rapid adoption. However, automated configurations achieved comparable performance in MIMIC-IV, suggesting that ontology enrichment may suffice for well-documented features. The retrieval scoring weights were optimized on the TNBC development set and may require adjustment for different documentation styles. The current pipeline captures entity co-occurrence rather than explicit semantic relationships; incorporating relation extraction models could enhance the extraction of relationship-dependent features. For TNBC, inter-rater agreement with an independent second annotator was not assessed. For categorical variables, we assumed the absence of documentation indicates the absence of a condition, which may introduce chart-verification bias [44]. The survival analysis is limited by a low event count (12 events), which restricts statistical power for equivalence testing; validation in larger cohorts with more events is needed to confirm that extraction accuracy translates to equivalent prognostic performance.

Future work will focus on improving retrieval strategies to address the primary source of errors and implementing ensemble approaches that route features to appropriately sized models based on complexity. We will expand validation to community oncology settings and pursue multi-institutional validation within the same cancer type to assess generalizability across EHR systems. Ultimately, we aim to integrate extracted phenotypes with imaging-derived features to enable multimodal outcome prediction at scale.

## 5. Conclusion

This study introduces OncoRAG, a graph-based RAG pipeline that extracts structured clinical features from unstructured oncology notes using a locally deployed mid-size language model without task-specific fine-tuning. Validated across two languages, three institutions, and two cancer types, the pipeline achieved F1 scores of 0.79–0.84 and yielded prognostic models with discrimination comparable to manual curation. By reducing the data extraction bottleneck with a locally deployable, computationally efficient approach, OncoRAG may enable larger-scale evidence generation from real-world oncology data.

## Data Availability

The code supporting the findings of this study, along with a simulated dataset for testing, will be made publicly available upon publication at https://github.com/pgsalome/oncorag. The institutional clinical data (TNBC and RiCi) cannot be made publicly available due to patient privacy, but may be accessible under appropriate institutional agreements.

## Supplementary Material

**Supplementary Table S1.**
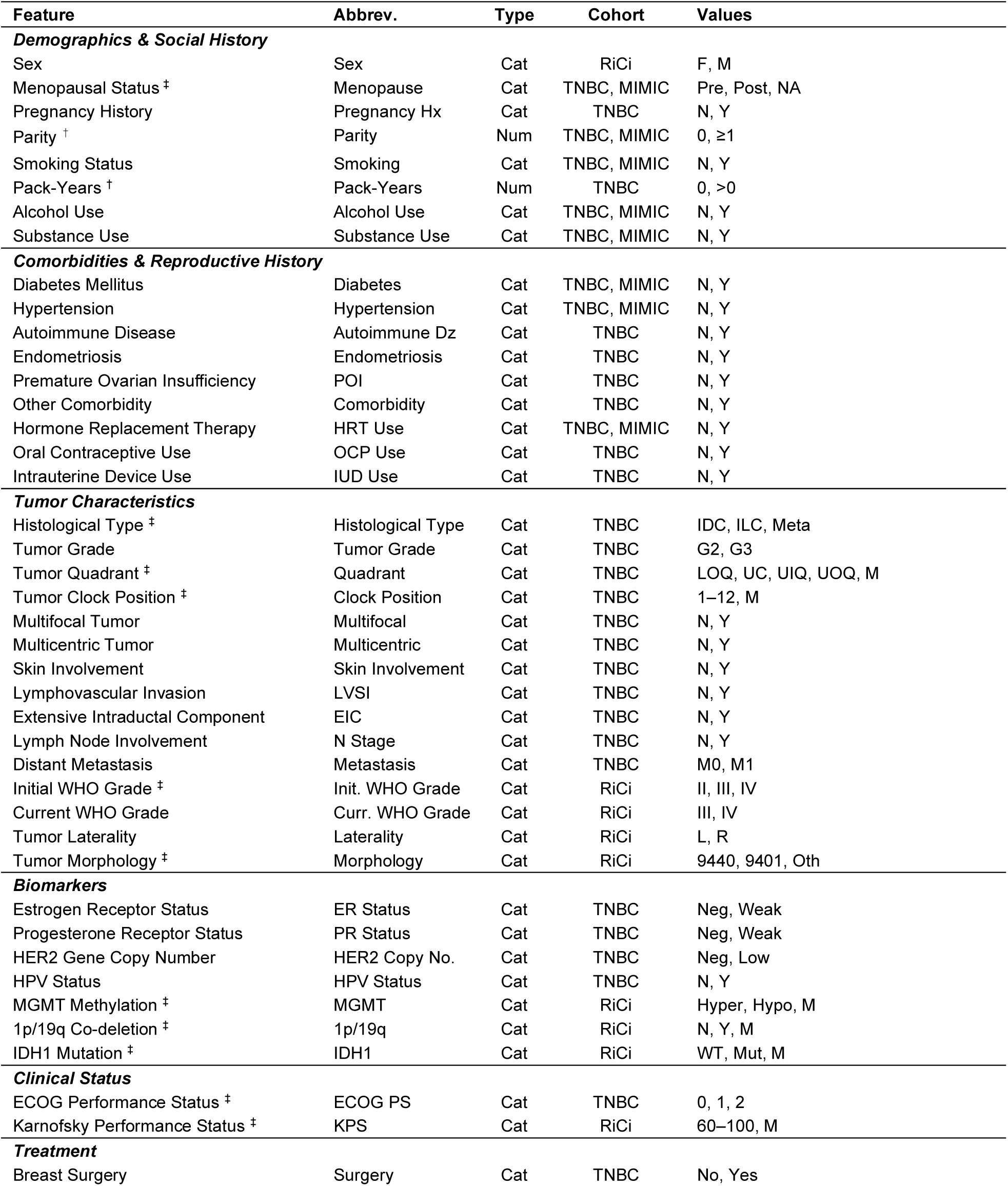
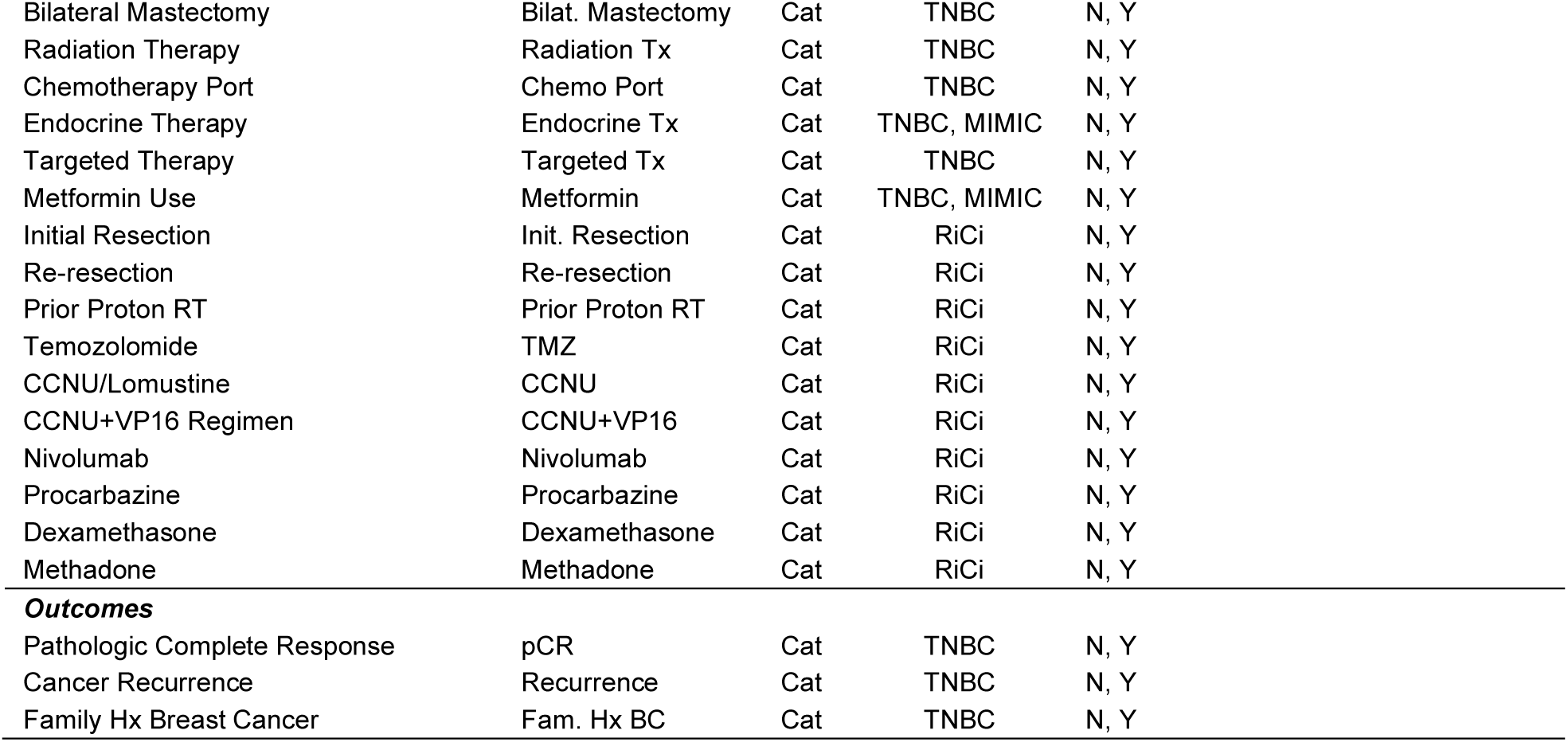
Feature definitions and evaluation categories. † originally continuous, binarized for evaluation; ‡ multiclass categorical (including features where missing is retained as a separate class); unmarked features are binary categorical. For Parity, evaluation was binarized in the TNBC cohort and multiclass (including NA) in MIMIC-IV. Cat, categorical; Num, numeric. Full abbreviations are defined at first use in the main text.

**Supplementary Table S2.** Knowledge graph node and edge types.

| Node Type | Key Attributes | Description |
| --- | --- | --- |
| Patient | patient_id | Root node |
| Note | note_id, file_name | Clinical document |
| Sentence | sentence_text, position | Source text containing entity mention |
| Entity | normalized_text, label, context flags | Extracted clinical entity |
| Date | date_string | Temporal reference |
| Edge Type | Connection | Description |
| has_note | Patient → Note | Links patient to documents |
| mentions | Sentence → Entity | Entity mentions with source sentence |
| contains | Note → Sentence | Links documents to their sentences |
| co_occurs | Entity ↔ Entity | Co-occurring entities (weighted) |
| has_date | Note → Date | Temporal reference |
| occurred_on | Entity → Date | Entity-date associations |

**Supplementary Table S3.** Example extraction prompt for BRCA1 mutation status, illustrating the four-component structure.

| Component | Content |
| --- | --- |
| 1. Task Definition | You are a specialized clinical data analyst extracting information from breast cancer medical records. Identify the patient's BRCA1 mutation status from clinical notes. |
| 2. Feature Description | The feature captures whether a BRCA1 mutation was detected, reported as negative, labeled as a variant of uncertain significance (VUS), or not reported at all. |
| 3. Rules and Examples | Ignore family history unless attributed to the patient. Distinguish orders or plans from confirmed results. Do not confuse BRCA2 mentions with BRCA1. If no explicit result is present, classify as Not Reported. Examples: "BRCA1 mutation detected" → Positive; "BRCA1 WT" → Negative; "BRCA1 VUS" → VUS; "Patient not tested" → Not Reported. |
| 4. Output Options | A = Positive; B = Negative; C = VUS; D = Not Reported |

**Supplementary Table S4.**
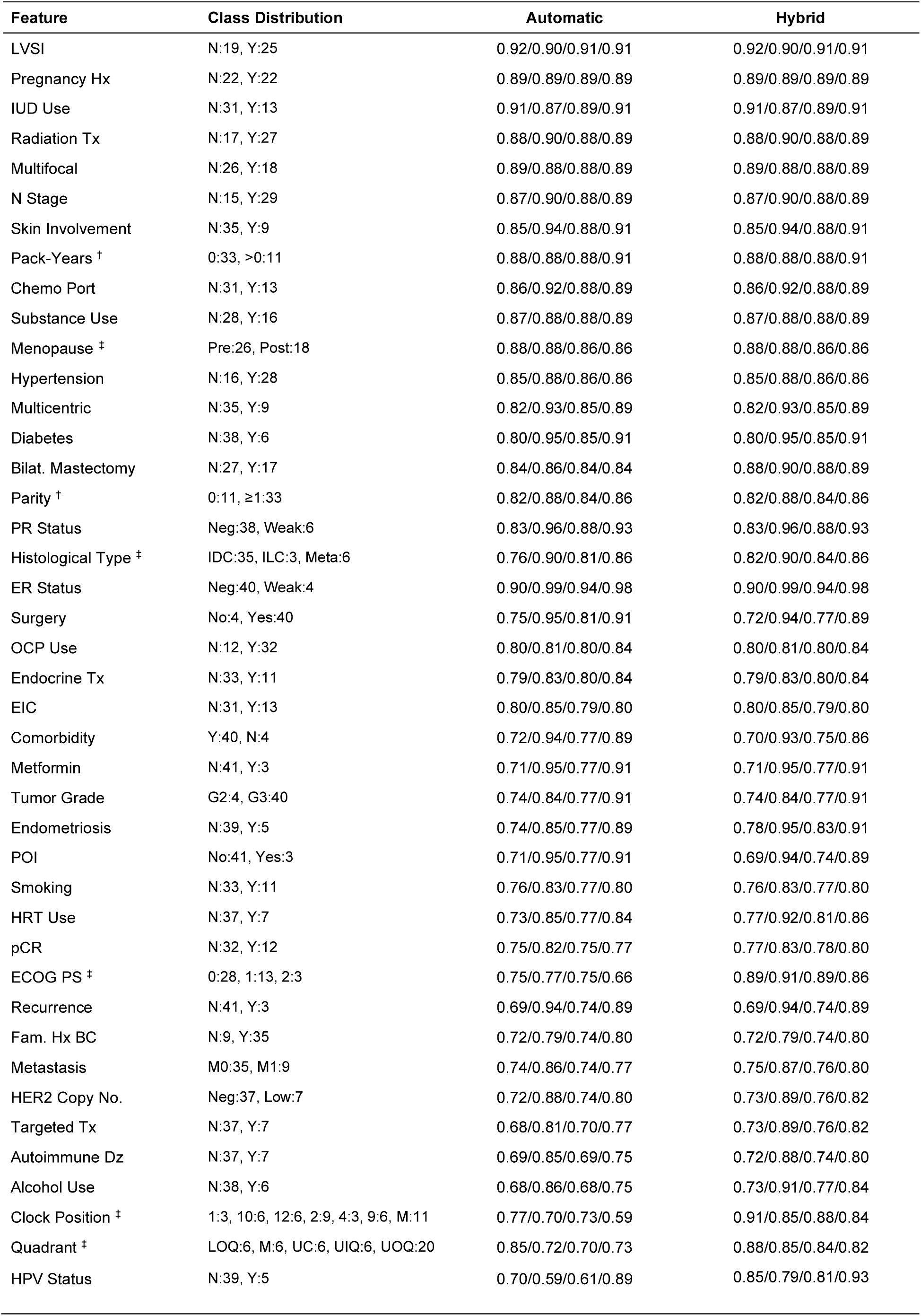
Feature-level extraction performance on the TNBC test set (n = 44 patients, 42 features). Values represent macro-averaged precision/recall/F1-score/accuracy. † binarized for evaluation; ‡ multiclass categorical. Ordered by automatic F1-score (descending).

**Supplementary Table S5.** Feature-level extraction performance on the RiCi test set (n = 61 patients, 19 features). Values represent macro-averaged precision/recall/F1-score/accuracy. ‡ multiclass categorical; M, missing (retained as a separate class). Ordered by automatic F1-score (descending).

| Feature | Class Distribution | Automatic | Hybrid |
| --- | --- | --- | --- |
| Methadone | N:58, Y:3 | 0.82/0.82/0.82/0.97 | 0.82/0.82/0.82/0.97 |
| Nivolumab | N:58, Y:3 | 0.88/0.99/0.92/0.98 | 0.88/0.99/0.92/0.98 |
| IDH1 ‡ | WT:33, Mut:2, M:26 | 0.95/0.96/0.95/0.93 | 0.96/0.95/0.95/0.93 |
| Procarbazine | N:59, Y:2 | 0.83/0.99/0.90/0.98 | 0.83/0.99/0.90/0.98 |
| Init. WHO Grade ‡ | II:2, III:4, IV:55 | 0.93/0.99/0.96/0.98 | 0.93/0.99/0.96/0.98 |
| Prior Proton RT | N:53, Y:8 | 0.83/0.96/0.88/0.93 | 0.90/0.98/0.93/0.97 |
| KPS ‡ | 90:12, 60:11, 80:8, 70:4, 100:3, M:23 | 0.95/0.88/0.87/0.85 | 0.95/0.88/0.87/0.85 |
| Dexamethasone | N:33, Y:28 | 0.88/0.86/0.87/0.87 | 0.87/0.87/0.87/0.87 |
| CCNU | N:39, Y:22 | 0.84/0.86/0.85/0.85 | 0.87/0.90/0.87/0.87 |
| CCNU+VP16 | N:57, Y:4 | 0.98/0.75/0.82/0.97 | 0.98/0.75/0.82/0.97 |
| Sex | F:23, M:38 | 0.75/0.77/0.75/0.75 | 0.93/0.93/0.93/0.93 |
| 1p/19q ‡ | N:4, Y:2, M:55 | 0.77/0.83/0.78/0.95 | 0.77/0.83/0.78/0.95 |
| Curr. WHO Grade | III:4, IV:57 | 0.72/0.96/0.78/0.92 | 0.97/0.74/0.82/0.96 |
| Morphology ‡ | 9440:55, 9401:2, Oth:4 | 0.69/0.97/0.77/0.92 | 0.77/0.83/0.79/0.97 |
| Laterality | L:33, R:28 | 0.68/0.67/0.65/0.66 | 0.84/0.79/0.79/0.80 |
| Init. Resection | N:11, Y:50 | 0.76/0.58/0.60/0.84 | 0.80/0.63/0.66/0.85 |
| Re-resection | N:33, Y:28 | 0.78/0.67/0.61/0.64 | 0.67/0.62/0.60/0.64 |
| TMZ | N:3, Y:58 | 0.57/0.75/0.58/0.82 | 0.57/0.75/0.58/0.82 |
| MGMT ‡ | Hyper:11, Hypo:14, M:36 | 0.57/0.58/0.56/0.66 | 0.57/0.58/0.56/0.66 |

**Supplementary Table S6.** Feature-level extraction performance on the MIMIC-IV test set (n = 100 patients, 10 features). Values represent macro-averaged precision/recall/F1-score/accuracy. † Binarized for evaluation; ‡ multiclass categorical; Ordered by automatic F1-score (descending).

| Feature | Class Distribution | Automatic | Hybrid |
| --- | --- | --- | --- |
| HRT Use | N:96, Y:4 | 0.87/0.87/0.87/0.98 | 0.87/0.87/0.87/0.98 |
| Menopause ‡ | NA:57, Pre:12, Post:31 | 0.83/0.82/0.82/0.85 | 0.83/0.82/0.82/0.85 |
| Parity † | M:57, 0:26, ≥1:17 | 0.95/0.95/0.95/0.97 | 0.95/0.95/0.95/0.97 |
| Substance Use | N:81, Y:19 | 0.91/0.93/0.92/0.95 | 0.91/0.93/0.92/0.95 |
| Diabetes | N:64, Y:36 | 0.86/0.83/0.84/0.86 | 0.86/0.83/0.84/0.86 |
| Metformin | N:89, Y:11 | 0.84/0.97/0.89/0.95 | 0.84/0.97/0.89/0.95 |
| Hypertension | N:32, Y:68 | 0.79/0.84/0.79/0.80 | 0.79/0.84/0.79/0.80 |
| Smoking | N:61, Y:39 | 0.80/0.81/0.80/0.81 | 0.80/0.81/0.80/0.81 |
| Alcohol Use | N:77, Y:23 | 0.90/0.75/0.80/0.88 | 0.90/0.75/0.80/0.88 |
| Endocrine Tx | N:98, Y:2 | 0.67/0.98/0.74/0.96 | 0.67/0.98/0.74/0.96 |

**Figure S1.**
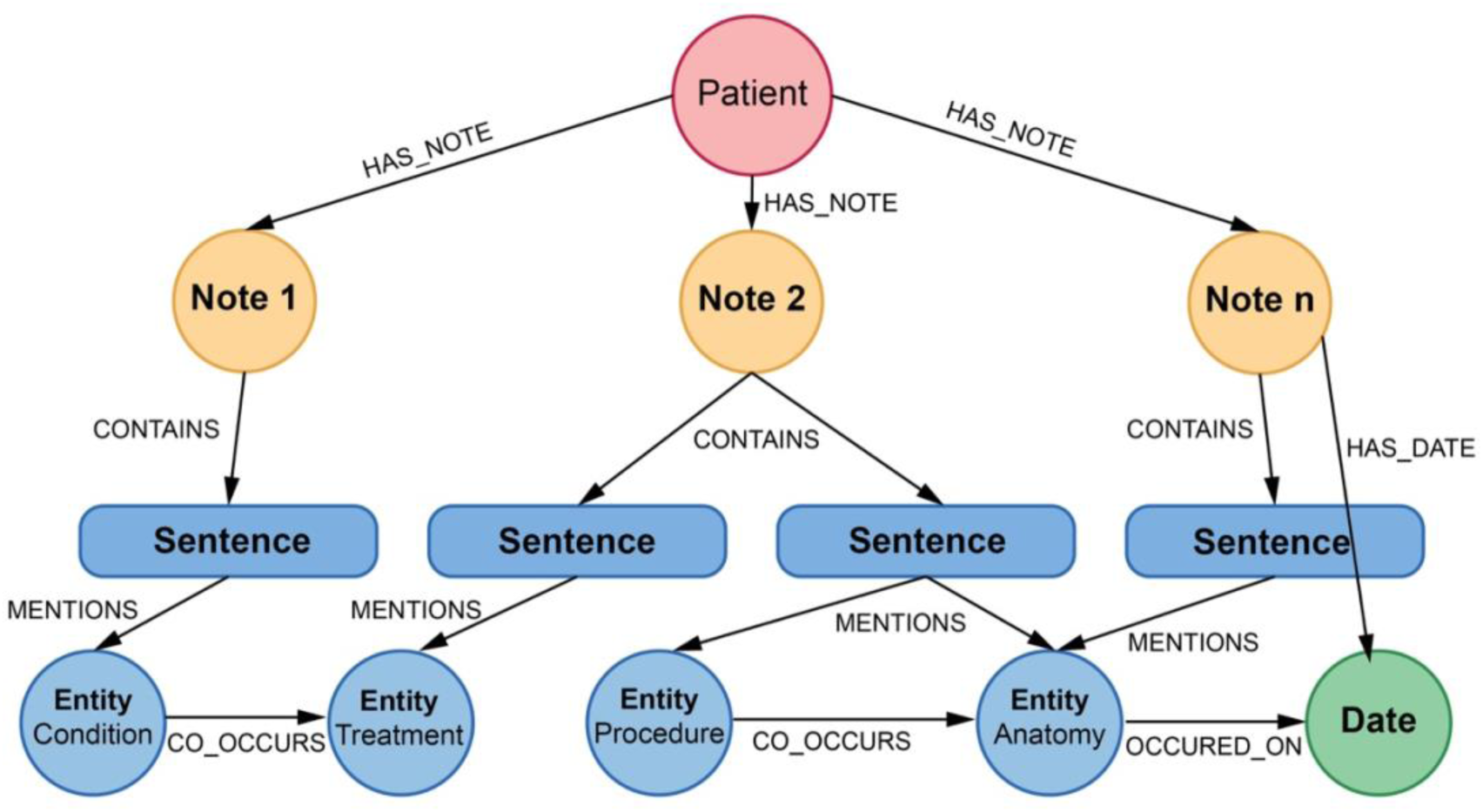
Clinical knowledge graph structure. Nodes represent patients, clinical notes, sentences, extracted entities (categorized as Condition, Treatment, Procedure, or Anatomy), and dates. Edges capture hierarchical relationships (has_note, contains), entity mentions within sentences (mentions), entity co-occurrences (co_occurs), and temporal associations (has_date, occurred_on). Sentence nodes preserve the source text for each entity mention, enabling context retrieval during feature extraction.

